# The mental health and experiences of discrimination of LGBTQ+ people during the COVID-19 pandemic: Initial findings from the Queerantine Study

**DOI:** 10.1101/2020.08.03.20167403

**Authors:** Dylan Kneale, Laia Bécares

## Abstract

**Objective:** To assess mental health status and experiences of discrimination amongst a sample of Lesbian, Gay, Bisexual, Transgender, Queer people (LGBTQ+, the “plus” including those who don’t identify with any such label) during the COVID-19 pandemic.

**Design:** Cross-sectional web-based survey.

**Setting:** Responses were collected during the COVID-19 pandemic between April 27^th^and July 13^th^.

**Participants:** 398 LGBTQ+ respondents forming an analytical sample of 310 in the main models.

**Methods:** We used a combined measure of gender identity or expression and sexual orientation as the main explanatory variable. We assessed mental health with the 4-item Perceived Stress Scale (PSS-4), and with the 10-item Center for Epidemiological Studies Depression scale (CES-D-10). We measured experiences of discrimination with a battery of questions that asked respondents whether they had experienced a set of discriminatory experiences because of their LGBTQ+ identity during the coronavirus pandemic. Experiences of discrimination was considered a mediating factor and examined both as an outcome as well as an explanatory variable. Models were adjusted for a range of demographic and socioeconomic variables.

**Results:** The prevalence of depression and stress were both high, with the majority of the sample exhibiting significant depressive symptomology (69%). Around one-in-six respondents reported some form of discrimination since the start of the pandemic because they were LGBTQ+ (16.7%). In regression models, the average score for perceived stress increased by 1.44 (95% Confidence Interval (CI): 0.517-2.354) for those who had experienced an instance of homophobic or transphobic harassment, compared to respondents who had not. Similarly, the odds of exhibiting significant depressive symptomology (CES-D-10 scores of 10 or more) increased three-fold among those who had experienced harassment based on their gender or sexuality compared to those who had not (OR: 3.251; 95% CI: 1.168-9.052). These marked associations remained after adjustment for a number of socioeconomic and demographic covariates. Cis-female respondents who identify as gay or lesbian had the lowest scores for perceived social or depressive symptoms; conversely transgender and gender diverse individuals had the highest scores.

**Conclusions:** We found high levels of stress and depressive symptoms, particularly among younger and transgender and gender diverse respondents. These associations were partially explained by experiences of discrimination which had a large, consistent and pernicious impact on stress and mental health.

## Introduction

The coronavirus pandemic has exposed and magnified existent societal and health inequities that operate across multiple and intersecting systems of oppression. Given documented stark health and socioeconomic inequalities across social locations related to sexuality, and gender expression and identity [1], the coronavirus disease (COVID-19) and subsequent social and economic implications could be expected to disproportionately impact on Lesbian, Gay, Bisexual, Transgender, and Queer (LGBTQ+, the “plus” including those who don’t identify with any such label) people. To date there is a dearth of information on whether this is the case.

Decades of relative empirical invisibility in health and social research means we have to draw from a limited evidence base in order to theorise the increased risks of exposure to COVID-19 and its impacts that LGBTQ+ people face. However, where evidence does exist, this overwhelmingly suggests that higher levels of pre-existing health conditions compared to cisgender and heterosexual populations, may place the LGBTQ+ community at additional risk of adverse prognosis. This includes long-term chronic illness, and higher rates of smoking and asthma among LGBTQ+ people [2-8]; higher rates of obesity, and alcohol consumption among lesbian, bisexual, and queer women [7 9 10]; and increased likelihood of being immunocompromised (e.g. HIV+ with a low CD4 cell count or with untreated HIV) among gay men and transgender people [11]. In addition, the impacts of social distancing and lockdown may be felt acutely by LGBTQ+ people, who even before the pandemic started, were at higher risk of poorer mental health as indicated by higher levels of suicide attempts and suicidal ideation, and lower levels of mental wellbeing [12 13].

Theoretical frameworks including the Minority Stress Model suggest that stark health inequalities are the result of distal and proximal stressors caused by living within ahomophobic, heterosexist, trans phobic culture, which often results in cumulative experiences of discrimination, harassment, victimization, expectations of rejection, and internalized transphobia and homophobia [14 15]. These experiences have been extensively documented across several (mostly US-based) studies, where high prevalence of experiences of stigma and discrimination has been reported among the LGBTQ+ community [16-20].

Within the heterogeneous LGBTQ+ umbrella term, individual groups may be positioned at a distinct disadvantage, for example with relation to gender expression and identity.Transgender and gender-diverse (TGGD) individuals have a current gender identity or expression that differs from the culturally-bound gender associated with one’s assigned birth [21 22]. TGGD identity is different from sexual orientation, which refers to the gender an individual is sexually or romantically attracted to. A growing body of evidence has shown that TGGD people experience higher rates of adverse mental health when compared to cis-gender individuals (people whose gender identity or expression matches their sex assigned at birth), particularly anxiety, depression, and suicidality [5 23-25]. They also report the highest prevalence of violence, marginalisation, and discrimination [16-19].

In the context of the current coronavirus pandemic, where existent social, economic, and health inequalities are being exacerbated, it is paramount to document whether, and how, inequalities between the LGBTQ+ community, and heterosexual, cis-gender individuals are being further amplified, given the health inequalities described above, and the lower socioeconomic status, and barriers to accessing services [26] experienced by the LGBTQ+ community.

The present study aims to address the limitations in the current evidence base by analysing data from the #Queerantine Study (a portmanteau Queer and Quarantine), understand web-based survey that assesses how LGBTQ+ people are experiencing the coronavirus pandemic, including the health, social, and economic impact of measures to reduce coronavirus transmission such as social distancing and the lockdown.

## Methods

Data collection was conducted via a cross-sectional, web-based anonymous survey using a structured questionnaire that began on 27 April 2020, and which is ongoing at the time of writing (August 2020). Data for the present analyses were collected until 13 July 2020. The target sample included respondents aged 18 and over, and who self-identify as lesbian, gay, bisexual, transgender, queer, as having another minority sexual orientation, gender non-binary, or as intersex. Cisgender respondents who self-identify as heterosexual were not excluded from the survey, although the recruitment and survey design were tailored towards measuring the experiences of the LGBTQ+ community. The #Queerantine survey asks respondents about their sociodemographic characteristics, their physical and mental health, health behaviours, and their experiences and anxieties relating to the pandemic and their identity. Input from a representative from the National LGB&T Partnership broadened the focus to consider how respondents had experienced changes in support from LGBTQ+ service providers and organisations; this input also helped to shape the measures around gender identity.

Ethical approval was obtained from the ethics board of the University of Sussex(ER/LB516/4) and University College London (REC 1335).

### Outcome variables

In this analysis we focus on two outcomes: mental health (including depression and stress), and experiences of discrimination. We assessed mental health with two validated measures, the 4-item Perceived Stress Scale (PSS-4), and the 10-item Center for Epidemiological Studies Depression Scale (CES-D-10). The PSS-4 assesses the extent to which situations in life are viewed as stressful [27]. The scale asks respondents about their ability to control important things in life, their confidence in handling personal problems, the extent to which respondents felt things were going their way, and whether difficulties were piling up so high they were becoming insurmountable, using the past month as a frame of reference. Scores range between 0 and 15, and have good levels of internal consistency in our analytical sample (Cronbach’s α=0.83).

The CES-D-10 asks respondents to consider how much in the past week they have experienced feelings including loneliness, happiness, and fear [28]. In total the scale includes three items on depressed affect, five items on somatic symptoms, and two on positive affect [29]. Scores range between 0 and 30, with thresholds used to denote ‘depressive symptoms’ based on a score of ten or over [28] (although this threshold may be slightly higher in some populations [29]). In the present study we examine the CES-D-10 both as continuous (Cronbach’s α=0.87), and as a binary measure, with a cut-off of 10 or more indicating significant depressive symptomology.

Experiences of discrimination were assessed with a set of questions that asked respondents whether, since the start of the coronavirus pandemic, they had experienced verbal harassment, physical harassment, sexual harassment, threats of violence, exclusion from events/activities, involuntary disclosure of LGBTQ+ identity, or other forms of inappropriate treatment because they were LGBTQ+ or were perceived as being LGBTQ+. Individual measures were combined into one summary variable of ‘any discrimination.’

### Sexual orientation and gender identity or expression

Gender identity was assessed using the recommended two-step method [30] with two items:(1) assigned sex at birth (female, male) and (2) current gender identity. The two items were cross-tabulated to categorise participants as either TGGD or cisgender.

Sexual orientation was captured with a question that asked participants to select their sexual orientation from the following categories: Bisexual; Gay/Lesbian; Heterosexual/Straight; Don’t know; Prefer not to say; and an Other, free-text category.

We examine sexual orientation and gender identity or expression as different constructs, and also combine both into a separate variable that examines the intersection of sexual orientation and gender identity or expression, using the following five categories: (i) cisgender female lesbian/gay; (2) cisgender female other non-heterosexual (including bisexual, other, don’t know, and prefer not to say); (3) transgender and gender diverse; (4) cisgender male gay; (5) cisgender male other non-heterosexual (including bisexual, other, don’t know, and prefer not to say).

### Covariates

We adjusted for variables thought to confound the association between sexual orientation, gender identity or expression, discrimination, and mental health. This included age, relationship status, ethnicity, and whether the respondent was based in the UK or not. We also controlled for socioeconomic status with a variable that asked respondents about their subjective social status (modelled on a validated approach [31]), and how this had changed since the start of the pandemic, with categories reflecting no change, positive change, and negative change.

### Analytical plan

The analysis consisted of a complete case analysis of respondents who provided data for the measures of interest; this included those who provided ambiguous responses including don’t know and ‘prefer not to say.’

Summary statistics were calculated for baseline characteristics overall and across the outcomes of interest. Associations between main exposure variables and outcomes were tested in unadjusted analyses using the χ^2^test of association and ANOVA as appropriate. Ordinary Least Squares regression models were constructed for continuous models of stress and depression, and binary logistic regression models were constructed for depressive symptomology. For models where mental health is the outcome of interest, the measure of discrimination and harassment was used as the main exposure variable.

For models where discrimination and harassment is the outcome of interest we only adjusted for sexual orientation, gender identity or expression, age, ethnicity, and location. An additional analysis examining when in the pandemic harassment and discrimination occurred is also included.

Adjusted and unadjusted models are presented. Analyses were conducted in Stata 14 [32].

## Results

Between April 27^th^and July 13^th^, a total of 426 responses were received. Of these 24 were excluded because they did not provide their age, and 4 were excluded because they were aged 18 or younger. Of the remaining 398, we were able to calculate PSS-4 and CES-D-10 scores for 325 and 324 respondents respectively. Once we had accounted for missingness on our other covariates, our analytical sample consisted of 310 respondents for models of mental health, excluding one further cisgender heterosexual respondent.

The analytical sample broadly mirrored the sociodemographic characteristics of the recent UK government National LGBT Survey [33]. The distribution of respondents by sexual orientation was very similar, albeit with a higher share of respondents who identified as Queer in the Queerantine survey (8% vs approximately 1%). The proportion of respondents aged 18-24 was lower at 15.1% (compared with approximately 37.4%), with higher proportions at older age groups in line with the UK population as a whole. Almost a quarter of the sample (23.5%) were categorised as TGGD, suggesting greater representation than in the National LGBT Survey sample where the proportion of TGGD respondents stood at approximately 15% [33].

Descriptively, the results suggested that the sample had high levels of stress and depression. The mean score for PSS-4 (Mean(M): 7.67; Standard Deviation (SD): 3.22) was higher than that observed in UK community samples in previous studies [34] and selected studies of sexual minorities conducted elsewhere [35].The mean scores for CES-D-10 were also high. Using the recommended threshold of 10 or more to identify depressive symptomatology, we observed high levels of respondents falling into this category (71.9%), a higher proportion than observed among other populations known to be susceptible to depression such as people living with HIV/AIDS [36].

Around one-in-six respondents reported some form of harassment since the start of the pandemic because they were LGBTQ+ (16.7%); the most common forms being verbal harassment including insults or other hurtful comments (8.7%), exclusion from events or activities (5.6%) and involuntary disclosure of LGBTQ+ identity (3.5%).

Bivariate analyses show that cis-female respondents who identify as gay or lesbian had the lowest scores for perceived stress or depressive symptoms (see Table 1); conversely transgender and gender diverse individuals had the highest scores, with the small number of cismale respondents who had another sexual orientation except for gay also having high stress scores.

**Table 1:** Mental health, experiences of discrimination, and sociodemographic characteristics of the Queerantine Study respondents.

|  | Cis female<br>Gay/Lesbian | Cis female<br>bisexual/<br>other/<br>don't<br>know/<br>prefer not<br>to say | Trans-<br>gender<br>and<br>Gender<br>Diverse<br>(TGGD) | Cis male<br>Gay | Cis male<br>bisexual/<br>other/do<br>n't<br>know/<br>prefer<br>not to<br>say | Total |
| --- | --- | --- | --- | --- | --- | --- |
|  | % | % | % | % | % | % |
| **Mental health outcomes** |  |  |  |  |  |  |
| PSS-4 Score,<br>M(SD) | 6.44<br>(3.18) | 8.33<br>(3.14) | 8.96<br>(2.99) | 7.03<br>(2.97) | 9.00<br>(3.37) | 7.672<br>(3.218) |
| CES-D-10,<br>M(SD) | 12.0<br>(6.65) | 15.0<br>(5.86) | 17.15<br>(6.6) | 12.75<br>(7.17) | 16.15<br>(7.5) | 14.174<br>(6.948) |
| **Depressive**<br>**Symptomology** |  |  |  |  |  |  |
| No evidence (<10) | 36.62 | 18.33 | 16.44 | 38.71 | 15.38 | 28.06 |
| Evidence of depressive<br>symptomology ($\geq 10$) | 63.38 | 81.67 | 83.56 | 61.29 | 84.62 | 71.94 |
| **Any harassment or**<br>**inappropriate incidents** |  |  |  |  |  |  |
| None reported | 74.65 | 81.67 | 64.38 | 87.10 | 84.62 | 77.81 |
| Harassment reported | 19.72 | 13.33 | 28.77 | 7.53 | 15.38 | 16.72 |
| No information | 5.63 | 5.00 | 6.85 | 5.38 | 0 | 5.47 |
| **Age Group** |  |  |  |  |  |  |
| 18-24 | 9.86 | 18.33 | 31.51 | 2.15 | 30.77 | 15.11 |
| 25-34 | 18.31 | 45.00 | 31.51 | 32.26 | 23.08 | 30.87 |
| 35-44 | 39.44 | 23.33 | 16.44 | 31.18 | 23.08 | 27.65 |
| 45-54 | 23.94 | 10.00 | 16.44 | 23.66 | 15.38 | 19.29 |
| 55+ | 8.45 | 3.33 | 4.11 | 10.75 | 7.69 | 7.07 |
| **Change in perceived social status** |  |  |  |  |  |  |
| Negative change in status | 25.35 | 26.67 | 36.99 | 23.66 | 23.08 | 27.65 |
| No change | 52.11 | 43.33 | 35.62 | 51.61 | 61.54 | 46.95 |
| Positive change | 22.54 | 30.00 | 27.40 | 24.73 | 15.38 | 25.40 |
| Total |  |  |  |  |  | 100.00 |
| **Relationship status** |  |  |  |  |  |  |
| Single | 21.13 | 25.00 | 42.47 | 19.35 | 23.08 | 26.37 |
| Dating or in a relationship but not living together | 21.13 | 30.00 | 19.18 | 26.88 | 46.15 | 25.08 |
| Cohabiting/Married/Civil Partnership | 56.34 | 45.00 | 34.25 | 49.46 | 30.77 | 45.98 |
| Divorced, Widowed or Prefer Not to Say | 1.41 | 0 | 4.11 | 4.30 | 0 | 2.57 |
| **Identify as ethnic minority** |  |  |  |  |  |  |
| Not an ethnic minority | 87.32 | 81.67 | 90.41 | 83.87 | 76.92 | 85.53 |
| Ethnic minority | 12.68 | 16.67 | 9.59 | 12.90 | 23.08 | 13.18 |
| Prefer Not to Say | 0 | 1.67 | 0 | 3.23 | 0 | 1.29 |
| **Location** |  |  |  |  |  |  |
| UK | 81.69 | 80.00 | 82.19 | 90.32 | 76.92 | 83.60 |
| Rest of the world | 18.31 | 20.00 | 17.81 | 9.68 | 23.08 | 16.40 |

Stress was markedly higher for those who had experienced discrimination (PSS-4 M: 9.44 SD: 2.99) compared to those who had not (PSS-4 M: 7.35 SD: 3.16). Respondents who had experienced discrimination also had higher depression symptomology scores (CES-D-10 M:17.87 SD: 6.21) compared to those who had not (CES-D-10 M: 13.43 SD: 6.97). ANOVA tests showed that the observed differences in both mental health outcomes were statistically significant for both exposure variables (p<0.001). Similar differences were observed using CES-D-10 scores of ten or over as a threshold for significant depressive symptoms, with over four-fifths of TGGD respondents and respondents who had experienced harassment showing evidence of significant depressive symptomology (83.6% and 90.4% respectively).

We examined the relationship between gender identity and sexual orientation and discrimination in logistic regression models (see Table 2). Based on the association observed in Table 1 where TGGD respondents appeared to be at highest risk of harassment, we used TGGD as the reference category and explored to what extent the higher risk of TGGD to experience discrimination remained after controlling for basic sociodemographic covariates. The results from adjusted models showed that the odds of experiencing discrimination were lower for all other groups, and significantly lower in the case of cisgender gay males (OR: 0.237, CI:0.091-0.617) and cisgender LGBTQ+ females who identified with a sexual minority orientation other than gay/lesbian (OR: 0.361, CI: 0.141-0.921). Within the sample, the results were suggestive of a u-shaped trend in terms of age, with the youngest and the oldest LGBTQ+ respondents in the sample being at greatest risk of experiencing discrimination, although differences by age were generally not statistically significant. Being of an ethnic minority background or being located outside the UK were not associated with experiences of discrimination among this sample.

**Table 2:**
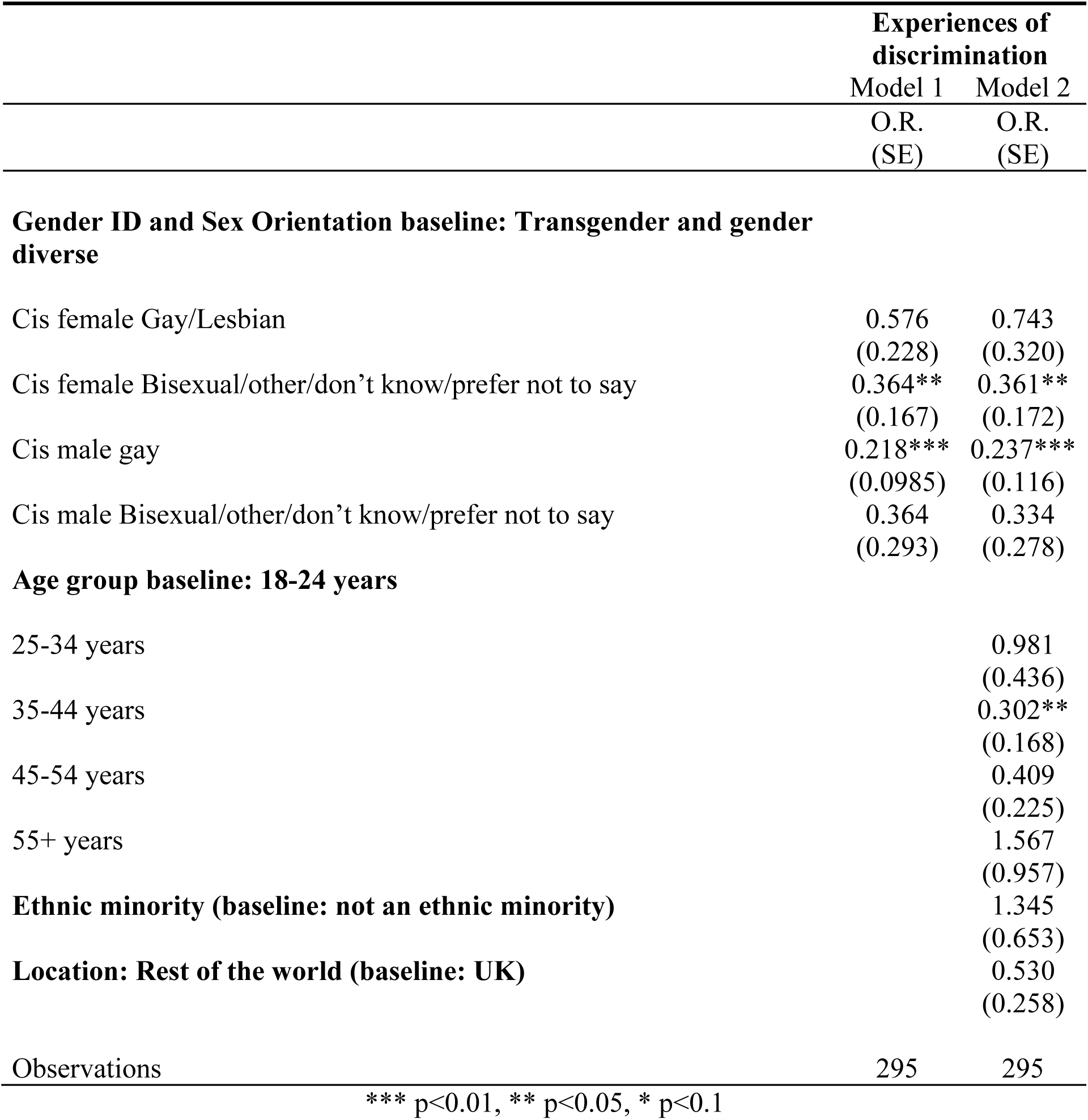
Logistic regression results for unadjusted and adjusted associations between gender identity/sexual orientation and discrimination during COVID-19 pandemic (Odds ratios and exponentiated standard errors in brackets)

We then moved to examine mental health, using discrimination as a key exposure variable alongside LGBTQ+ category (see Table 3). Experiences of discrimination were clear predictors of poorer mental health status among our LGBTQ+ sample. The average score for perceived stress increased by 1.44 points (95% Confidence Interval (CI): 0.517-2.354) for those who had experienced discrimination, compared to those who had not. Similarly, the odds of exhibiting significant depressive symptomology (CES-D-10 scores of 10 or more) increased three-fold among those who had experienced discrimination based on their gender or sexuality compared to those who had not (OR: 3.251; 95% CI: 1.168-9.052). These marked associations remained after adjustment for a number of covariates (see Models 1 to 4 in Table 3).

**Table 3:**
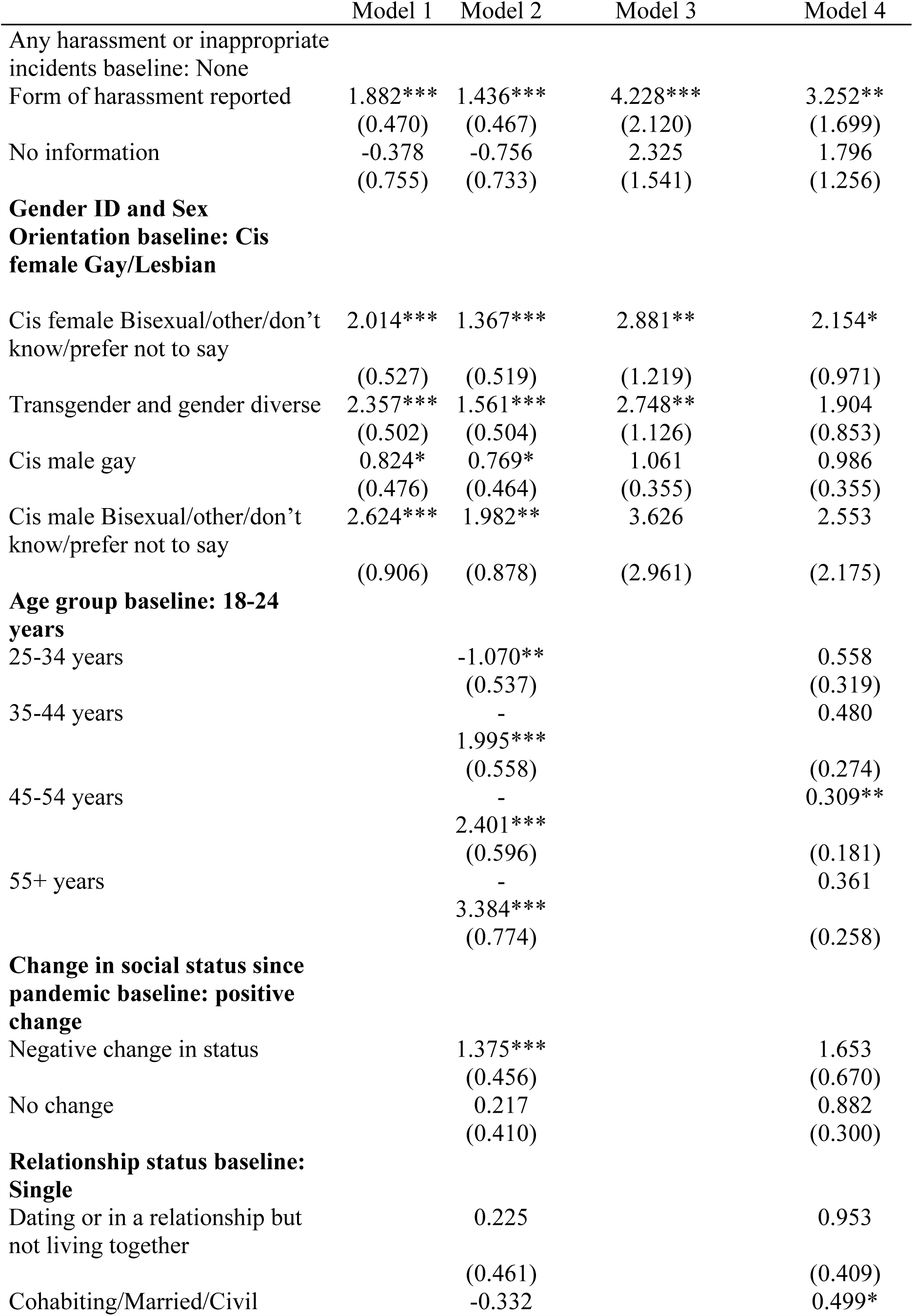
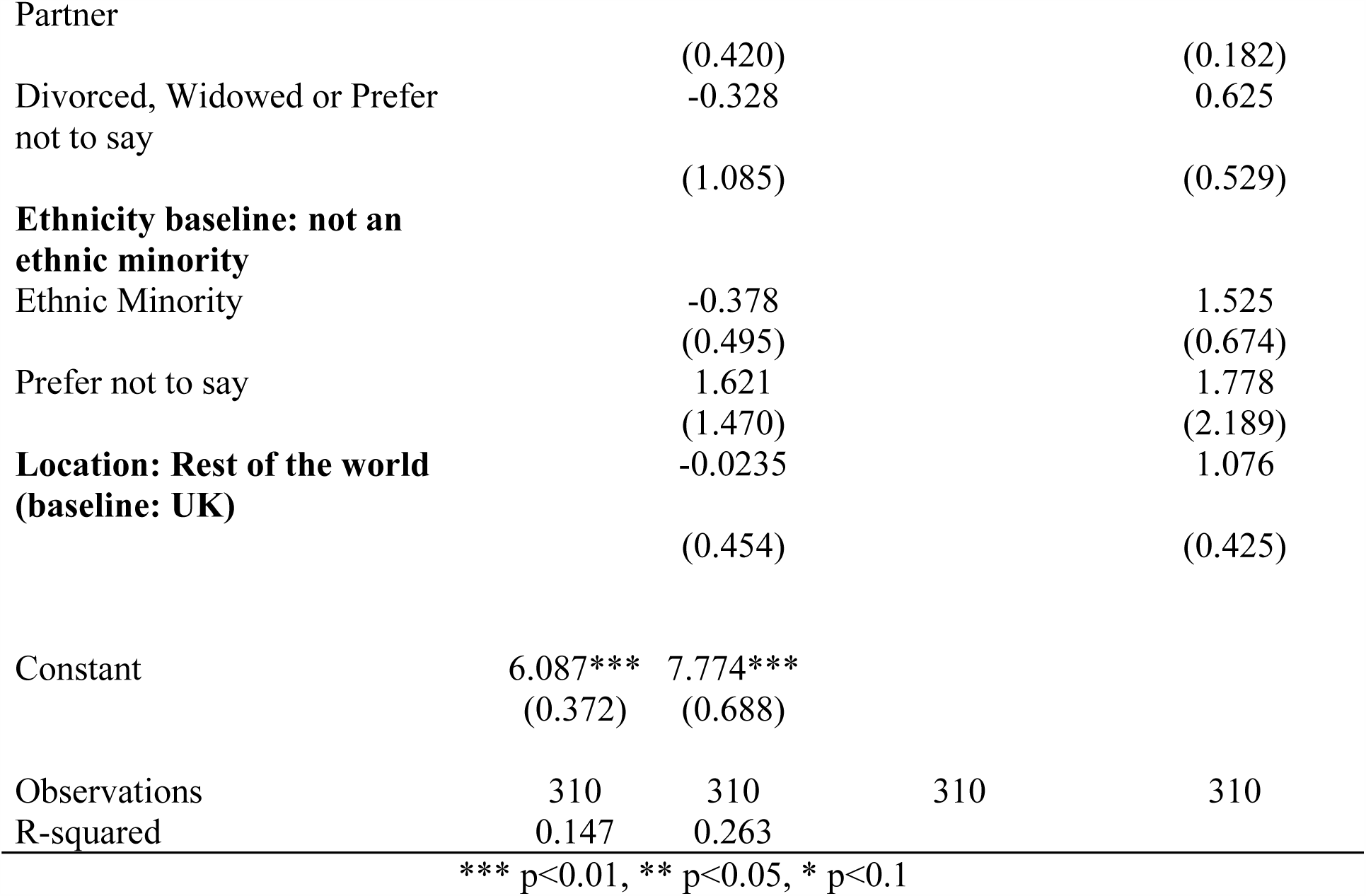
Results of unadjusted and adjusted OLS Regression for PSS-4 score (Models 1 and 2; regression coefficients and standard errors in brackets) and unadjusted and adjusted logistic regression results for odds of significant depressive symptomology indicated by CES-D-10 scores ≥10 (Models 3 and 4; odds ratios and exponentiated standard errors in brackets)

Cis female lesbian or gay respondents had better mental health than other LGBTQ+ groups, with average stress scores being statistically significantly higher for every other LGBTQ+ category. TGGD respondents and non-heterosexual cisgender males who did identify as gay had among the highest average stress scores after adjusting for other covariates. Although a similar trend was observed in the odds of experiencing significant depressive symptoms, this did not reach statistical significance. Given the high levels of significant depressive symptoms in the sample, we may conclude that a threshold of 10 is simply inadequate in discriminating those with the most severe depressive symptomology, given the high scores and large proportions of sample members falling within this threshold. Further exploration of the CES-D-10 as a continuous measure (see appendix table) shows that TGGD had statistically significantly higher CES-D-10 scores than cis gender lesbian or gay females, with an average score 3.38 points higher after adjusting for other covariates.

A clear trend by age was observed in models 3 and 4 (see Table 3), with younger respondents having significantly poorer mental health than those individuals in older age groups, both for stress and significant depressive symptomology, after adjusting for other covariates. The results show unambiguously that younger LGBTQ+ people in the sample had markedly higher levels of stress and depressive symptoms during the pandemic than older LGBTQ+ people.

## Discussion

In this study we present data on the mental health and experiences of discrimination of LGBTQ+ people during the COVID-19 pandemic. We show that scores for validated measures of stress and depressive symptoms among our LGBTQ+ sample are high, and higher than observed in community samples and vulnerable populations in the recent past (for example [34 36]). Furthermore, the pandemic may not be impacting the LGBTQ+ acronym evenly, with TGGD individuals having particularly high scores for stress and depressive symptoms relative to cisgender gay and lesbian individuals. Non-heterosexual respondents who are cisgender but do not identify as lesbian or gay also had elevated scores for stress and depressive symptoms. Similarly, there was a clear age gradient with younger LGBTQ+ people having much higher risks of showing symptoms of stress and depression.

Our analyses of discrimination reinforce the theoretical basis for undertaking analyses of LGBTQ+ health and mental health, with LGBTQ+ people theorised at greater risk of health issues due to a unique set of internal and external homophobic, heteronormative, and transphobic stressors [37]. We found that almost one-in-five respondents reported experiencing some form of discrimination during the pandemic, with TGGD respondents again at heightened risk of experiencing discrimination relative to other LGBTQ+ groups. Our results show that experience of discrimination was a mediating factor for higher stress and depressive symptomology, and the odds of individuals who had experienced discrimination also having significant depressive symptomology were three times higher than among individuals who had not experienced any discrimination. Open ended responses to the survey described various experiences of discrimination and inappropriate incidents including increased or excessive scrutiny, misgendering, and online abuse.

To further understand the results, we explored how mental health and discrimination varied over the course of the pandemic. We observed that mental health scores in the sample were poorer during the period April 27^th^-May 10^th^(the moment of ‘maximum risk’ as defined by the UK Prime Minister [38]) and during the period between May 23^rd^-June 14^th^(coinciding with revelations of lockdown breaches in the UK, transphobic comments on social media made by high profile people, and protests surrounding the shooting of George Floyd), although these differences were not significant. Similarly, we observed non-statistically significant differences in the proportion of respondents reporting instances of discrimination, with the initial easing of the lockdown and particularly the period from June 15^th^onwards coinciding with increases in harassment (see Figure 1), albeit based on a small sample in the latter period.

**Figure 1:**
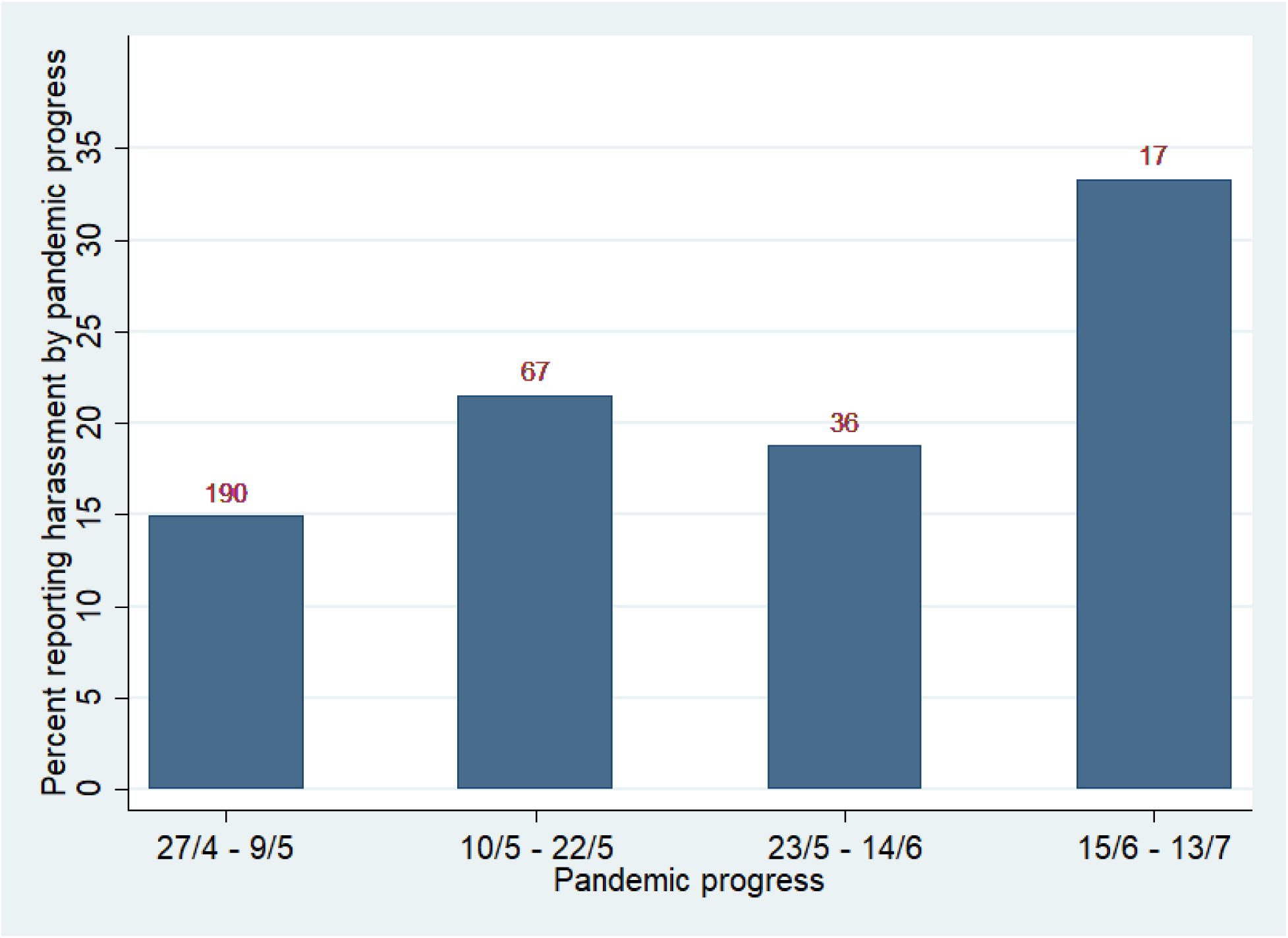
Proportion of respondents reporting discrimination by period in the pandemic.

## Limitations

Due to relatively small sample sizes, we have not been able to fully examine the diversity of the LGBTQ+ community, and fully examine how experiences vary according to social locations such as ethnicity, age, and gender identity (i.e., non-binary transgender people, non-heterosexual TGGD people, or ethnic minority lesbian women). Studies in the US show that the highest levels of violence are reported among transgender women of colour, and among young and low-income transgender people [19 39], suggesting that violence on the basis of transgender identity or expression often affects the most marginalized transgender subpopulations. Although we have adjusted for these factors in our models, we have not been able to further disaggregate across social locations to examine the role of interlocking systems of oppression in patterning experiences of discrimination and adverse mental health.While our data collection efforts are limited by the inherent challenge of surveying a small, dispersed, diverse, and difficult to reach population, it is nonetheless critically important to study the lives and experiences of discrimination and mental health among LGBTQ+ communities because of the stark health and social inequalities they experience.

New purposeful data collection was deemed appropriate as although a number of large representative studies (e.g. the UK Household Longitudinal Study) are currently collecting data on COVID-19 experiences, they typically contain small numbers of LGBTQ+ people [40 41], often do not collect information on TGGD identities, and contain heteronormative measures that can be exclusionary to LGBTQ+ respondents. An online convenience sample was deemed appropriate due to the absence of robust data on LGBTQ+ people from large surveys that could help to determine the characteristics of a representative sample of LGBTQ+ people. An online approach was particularly suitable for those respondents who may have been sheltering or shielding in households where their LGBTQ+ status was unknown to other members of the household [42]. Furthermore, this approach is in line with other recent large scale efforts at understanding the health of LGBT people in the UK [33]. We do, nevertheless, acknowledge that an online convenience sample can introduce potential issues around sample selection and the possibility that those living in stressful situations or with depressive symptoms were more likely to self-select into the survey.

### Implications of the findings

Results from the Queerantine Study suggest that groups within LGBTQ+ acronym may be at differential risk of exhibiting stress or depressive symptomology, although the sample as a whole may also be at higher risk than the general population of stress and depressive symptomology due to minority stress. The impact of homophobic and transphobic harassment and exclusion experienced during the pandemic has a deleterious impact on LGBTQ+ mentalhealth, demonstrated by the strong and consistent associations between harassment and poorer mental health in the models. Open-ended responses to survey questions emphasise the importance of LGBTQ+ social networks, often facilitated by the work of LGBTQ+ organisations, in providing support to LGBTQ+ individuals. However, these are the very organisations who are facing financial challenges with many now on the brink of closure [43]. There is very limited data at time of writing about the impact of COVID-19 on the broader LGBTQ+ community, and on TGGD people specifically, with most evidence coming from reports of voluntary, community, and social enterprise (VCSE) organisations. These reports highlight disproportionate impacts of the coronavirus pandemic on the LGBTQ+ community, and show that TGGD people report an increase in domestic violence and difficulties accessing sexual health services and gender-affirming medical treatment [44 45], which is thought to pose a particular risk for the mental health of transgender people awaiting such procedures [46].

Globally, LGBT+ rights organisations have alerted governments and public health officials about the need to address the vulnerability of the LGBTQ+ community to the coronavirus pandemic, including collecting sexual orientation and gender identity data for COVID-19 cases, increased socioeconomic support for disadvantaged individuals, and support for organisations working with the community [47]. Our findings give support to these demands given the documented high prevalence of depression and stress, and the concerning reports of experiences of discrimination. Poor LGBTQ+ mental health may remain unchecked without a substantial policy commitment and funding directed to ameliorating health inequalities exacerbated by the pandemic.

## Data Availability

Data are not available at the moment.

## Acknowledgments

We would like to thank the participants of the #Queerantine Study for sharing their experiences with us. We would also like to thank Harri Weeks and the National LGB&T Partnership for their advice and to all those who have helped to promote the study through social media and in other ways.

Despite our best efforts, this study was not funded by the UKRI (ESRC) COVID-19 Research initiative. At time of writing £0 (out of £90,491,960) had been awarded by the UK Research Councils to projects studying the experiences of the LGBTQ+ community during the coronavirus pandemic.

